# Cerina- Cognitive Behavioural Therapy based mobile application for managing GAD symptoms among Ulster University Students in Northern Ireland: A Protocol for a Pilot Feasibility Randomized Controlled Trial

**DOI:** 10.1101/2024.05.04.24306859

**Authors:** O. Eylem-van Bergeijk, S. Poulter, K. Ashcroft, T. Robinson, P. Mane, M. Islam, J. Condell, G. Leavey

## Abstract

**Introduction:** University students are one of the most vulnerable populations for anxiety disorders worldwide. In Northern Ireland, anxiety disorders appear to be more common among the university student population due to the population demographics across the region. Despite the need, these students show less inclination to access the widely available on-campus well-being services and other external professional services. Digital Cognitive Behaviour Therapy (CBT) aims to bridge this gap between the need for psychological help and access to it. However, challenges such as limited reach, low adoption, implementation barriers and poor long-term maintenance are mainstay issues resulting in reduced uptake of digital CBT. As a result, the potential impact of digital CBT is currently restricted. The proposed intervention “Cerina” is a scalable CBT-based mobile app with an interactive user interface that can be implemented in University settings if found to be feasible and effective.

**Methods and analysis:** The study is a single-blind pilot feasibility Randomized Controlled Trial (RCT) aiming to test the feasibility and preliminary effects of Cerina in reducing Generalised Anxiety Disorder (GAD) symptoms. Participants are 90 Ulster University students aged 18 and above with self-reported GAD symptoms. They will be allocated to two conditions: Treatment (i.e., access to Cerina for 6 weeks) and a waitlist control group (i.e., optional on-campus wellbeing services for 6 weeks). Participants in the waitlist will access Cerina 6 weeks after their randomization and participants in both conditions will be assessed at baseline, at 3 (mid-assessment), and 6 weeks (post-assessment). The primary outcome is the feasibility of Cerina (i.e. adherence to the intervention, its usability and the potential to deliver a full trial in the future). The secondary outcomes include generalised anxiety, depression, worry and quality of life. Additionally, participants in both conditions will be invited to semi-structured interviews for process evaluation.

**Ethics and dissemination:** Ethical approval for the study has been granted by the Ulster University Research Ethics Committee (ID: FCPSY-22-084). The results of the study will be disseminated through publications in scientific articles and presentations at relevant conferences and/or public events.

**Trial registration:** ClinicalTrials.gov ID NCT06146530

**Strengths and Limitations:**

- Low-threshold CBT-based mobile application, easily accessible to university student population
- Partnership between Ulster University, Ulster Student Wellbeing team and a mental health start-up to increase the reach and access the target population and evaluate the intervention.
- The study is designed as a feasibility trial and is not powered to detect statistically significant effect of the intervention.
- The study results might not generalise beyond the inclusion and exclusion criteria of the current study.
- The waitlist control groups are known to overestimate the effects of the intervention compared to treatment as usual

## Introduction

It is estimated that 264 million adults around the globe have anxiety (1). Of these adults, 179 million are female (63%) and 105 million are male (37%) (2). Anxiety disorders are very costly and are one of the leading contributors to the global health-related burden (3). According to the data on anxiety disorders obtained from the World Health Organization (WHO) Global Burden of Disease Study in 2019, the relative risk of the incidence of anxiety disorders and the disability-adjusted life years increased with age in adolescence and middle age and then decreased among people of both sexes (4). The most common anxiety disorder is Generalised Anxiety Disorder (GAD) affecting almost 30-40% of the global population (1). It is estimated that the prevalence of GAD varies widely across countries, with lifetime prevalence highest in high-income countries (5.0% [0.1%]), lower in middle-income countries (2.8% [0.1%]), and lowest in low-income countries (1.6% [0.1%]) (2). Lifetime comorbidity is high (81.9% [0.7%]), particularly with mood (63.0% [0.9%]) and other anxiety (51.7% [0.9%]) disorders. Treatment is sought by approximately half of the affected individuals (49.2% [1.2%]) in high-income countries (3). This means that GAD is untreated and unrecognised (5).

University students are one of the vulnerable populations for common mental health conditions (CMDs) in general and anxiety disorders specifically (6–8). According to the WHO, one out of three students was diagnosable with at least one CMDs, the most prevalent anxiety disorder in 2018 (6). During the COVID 19 pandemic, anxiety disorders alone affected about one in three students (9–11). The existing longitudinal studies do not consistently support the increased risk for GAD among students in the long term (12–14). However, they indicate a potential sub-group of students such as those who report more financial uncertainty or a history of CMDs, who could be more at risk of severe GAD and co-morbid CMDs and problems (13). From the life course perspective (15), university years correspond to emerging adulthood that is characterised by a transition period between adolescence and adulthood (16). On the one hand, ongoing life transitions and developmental alterations in this period can offer opportunities for self-exploration and growth (17). While on the other hand, such life transitions can exacerbate uncertainty, ambivalent feelings and risky behaviours (15). Additionally, international students and students from ethnic and sexual minority backgrounds may experience additional stressors related to culture shock, discrimination and lack of social and cultural capital (18). These stressors can interact with pre-existing vulnerabilities and well-known stressors such as academic pressure, financial uncertainties due to large amounts of study loans, and can lead to heightened stress which is central to the onset of physical and psychological conditions (19,20).

In Northern Ireland (NI), mental health problems and anxiety disorders are especially prevalent among university students due to the population demographics across the region in addition to the above mentioned factors common to the global university student population (9, 21,22). According to the data collected at Ulster University as part of the WHO World Mental Health International College Student Project (WMH-ICS), receipt of treatment was low (37.8%) (N=392), particularly among males and those with life time mental health condition with no suicidal thoughts and behaviours (STBs) (23). Males were less inclined to seek external professional services. However, interestingly they were less likely than females to rate embarrassment (OR=0.60) and/or worry about being treated differently (OR=0.63) as reasons for not seeking help (23). Additionally, those with a life-time mental health condition reported lower perceived need and/or readiness to change, and they rated being unsure about where to go as a more important barrier (OR=1.80) compared to those with STBs and without a life-time mental health condition (23). Moreover, a recent study conducted at Ulster University based on WMH-ICS data indicated that 22.6% (N=739) of students reported GAD symptoms, and students older than 21, those from LGBTQ+, and low SES backgrounds were less likely to access available psychological help (9). These results highlight vulnerability in line with the literature on mental health problems and the barriers to help-seeking among university student populations (24,25). Therefore, early interventions addressing the need for psychological care and treatment of diverse student populations are crucial even if they do not have suicidal thoughts (23).

Cognitive Behavioural Therapy (CBT) and pharmacotherapy are recommended evidence-based first-line interventions for GAD (26). CBT is equally effective as pharmacotherapy at the short term, but has larger overall treatment effects and fewer side effects than pharmacotherapy at the longer term (6-12 months follow-up) (g=0.34) (27). Despite the effectiveness and the favourable benefits of CBT over medication, there are recognised shortcomings in the current treatment of GAD resulting in a lengthy delay in the help-seeking process (6). Face-to-face CBT may not be accessible to everyone in need due to social (e.g. perceived stigma), organisational (e.g. waiting times and service availability), or financial reasons (e.g. in healthcare contexts where one must pay for their treatments) (28,29).

Digital CBT including CBT-based smartphone applications and web-based interventions may help overcome the above-mentioned barriers. They allow anonymity and accessibility. Also, they can reduce the costs associated with the provision of human resources by providing low-threshold access to evidence-based treatment among those who are in need (30). The existing literature point to mixed results about the effectiveness of these interventions in reducing GAD symptoms among student populations. For instance, a Randomized Controlled Trial (RCT) investigating the effectiveness of a guided, individually tailored web-based CBT programme, ICare Prevent in the Netherlands, indicated no evidence of a difference between the intervention and treatment as usual in anxiety and any of the examined outcomes (31). However, other RCTs investigating the same intervention reported promising results in terms of the feasibility, acceptability, and potential effectiveness of ICare Prevent among students with anxiety and/or depression in Germany (32,33) in South Africa (34) and Indonesia (35). Further support for the feasibility and potential effectiveness of digital CBT interventions among students have been provided by other RCTs (36,37).

Despite the obvious benefits of digital CBT, engaging with the intended users can be challenging. The percentage of those who complete the sessions ranges from 10% to 76% and dropout ranges from 30% to 40%. (38,39–41). Furthermore, an open feasibility trial by Lattie and colleagues (42) indicated that only a relatively small portion of a sample of university students (26.5%; 31/117) downloaded one or more of the CBT-based apps tested in the study, and approximately 24% (28/117) participants used these apps only once (42). Recent systematic reviews pointed out that two intervention methods show promise in mitigating the problem of low engagement with digital interventions (43,44). One approach aims to improve engagement through personalised feedback, customised notifications, and reminders (43). An alternative approach is aimed at changing users’ attitudes or behaviours by incorporating game-like strategies such as avatars (i.e. virtual therapists) or chatbots initiating conversations with users about specific content and/or exercises (43,44).

Overall, there are mixed results for the effectiveness of digital CBT interventions, and their impact is restricted by the issues related to limited reach, low adoption, implementation barriers and poor long-term maintenance among the intended users (45). Nevertheless, there is a need to test further approaches to address generalised anxiety symptoms among university student populations. Furthermore, there is a significant scope to improve the adherence to digital CBT-based interventions based on the above-mentioned intervention approaches utilising emerging technologies. In light of this, Cerina, a mental health start-up based in the UK, has developed a CBT-based mobile application for treating GAD symptoms which aims to address the unmet need through an interactive User Interface (UI) incorporating game-like strategies such as a chatbot, and engagement features such as customised notifications and pop-up reminders. The aims of the proposed study are two-fold: 1) To learn about the feasibility (i.e. adherence to the intervention, its usability and potential to deliver a full trial in the future) of Cerina among a university student population; and 2) to test the preliminary effects of Cerina in reducing GAD symptoms compared to a wait-list control group among Ulster University students presenting with self-reported GAD symptoms.

## Methods

### Setting

The study will be carried out in partnership between Cerina, a company developing CBT-based mobile applications for treating psychological disorders, and Ulster University as part of the NWE INTERREG IT4Anxiety project. The overarching aim of the IT4Anxiety project is to support the implementation of innovative solutions through start-ups to reduce anxiety. Ulster University has approximately 30,000 students enrolled in undergraduate, post-graduate and e-learning programmes across its four main campuses: Belfast, Coleraine, Derry Londonderry (Magee) and Jordanstown in Northern Ireland.

### Study Design

The study is a single-blind pilot feasibility Randomized Controlled Trial (RCT) with two conditions: Treatment and a Waitlist control group.

Eligible participants who have completed the baseline questionnaire will be randomized either into the treatment group in which they have access to the Cerina application for six weeks or into the wait-list control group in which they have access to the on-campus wellbeing services until the intervention group completes Cerina. They will then receive access to Cerina. Please see table 1 for the study design and figure 1 for the participant flow through trial.

**Figure 1.**
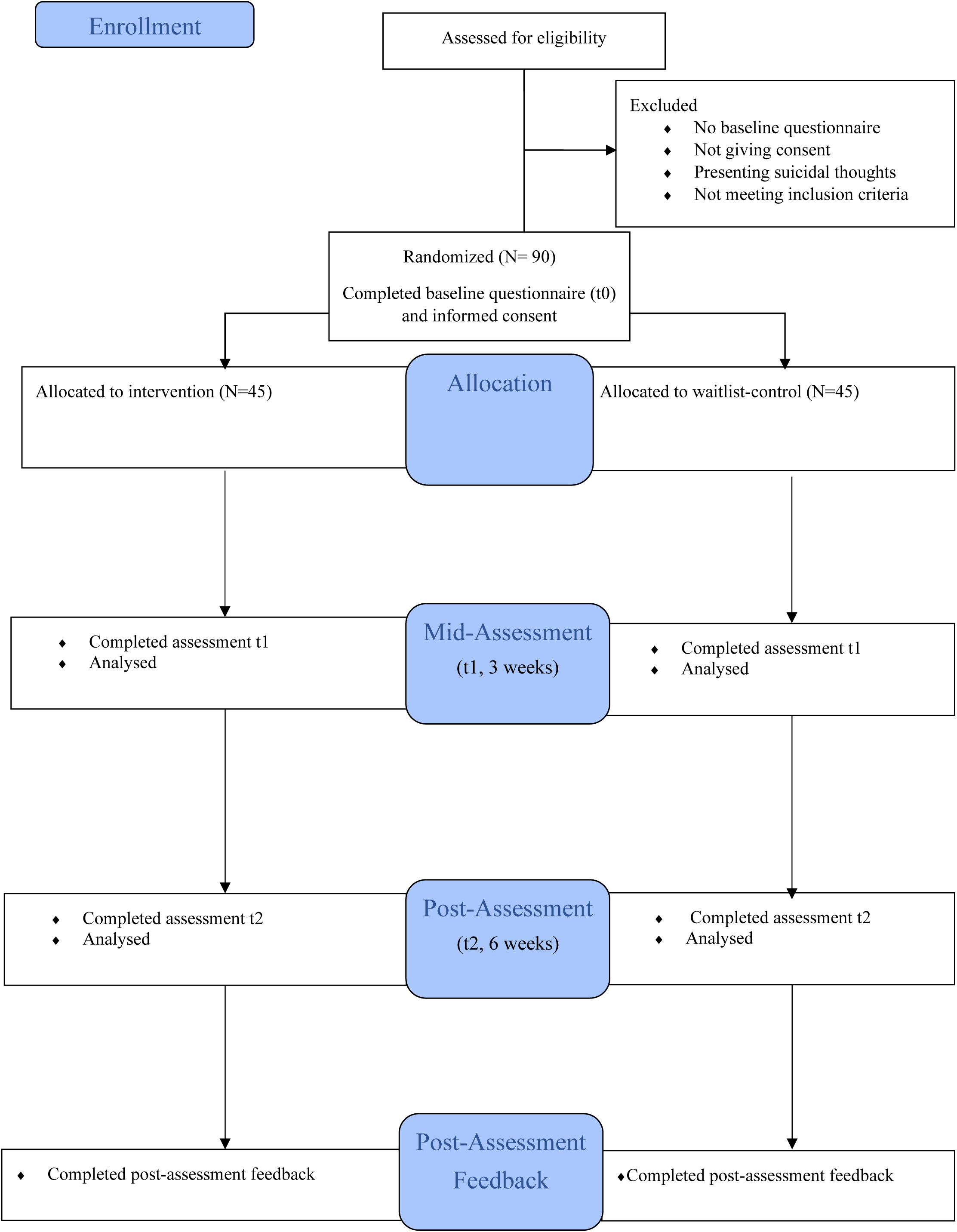
Participant flow through trial

**Table 1.** Study Design.

| Condition | Pre-test<br>T0 |  | 3 weeks<br>T1 | 6 weeks<br>follow-up<br>T2 |  |
| --- | --- | --- | --- | --- | --- |
| Treatment | X | Cerina | X | X |  |
| Wait-list Control | X | Routine<br>Care | X | X | Cerina |

All participants will be asked to fill in online self-report measures of GAD, depression, worry, and work and social adjustment at baseline (t0), at 3 weeks (t1), and at 6 weeks (t2) (see Table 2). Additionally, semi-structured interviews will be conducted with the participants to explore which elements of the intervention facilitated or hindered participants’ progress during the study and how the intervention can be optimised to increase its acceptability, relevance, and user-friendliness in real-life settings. Intervention group participants will be invited to the semi-structured interviews right after they complete their follow-up assessments at week 6. Wait-list control group participants will also be invited to the interviews after they access Cerina and complete their 6-week follow-up assessments.

**Table 2.**
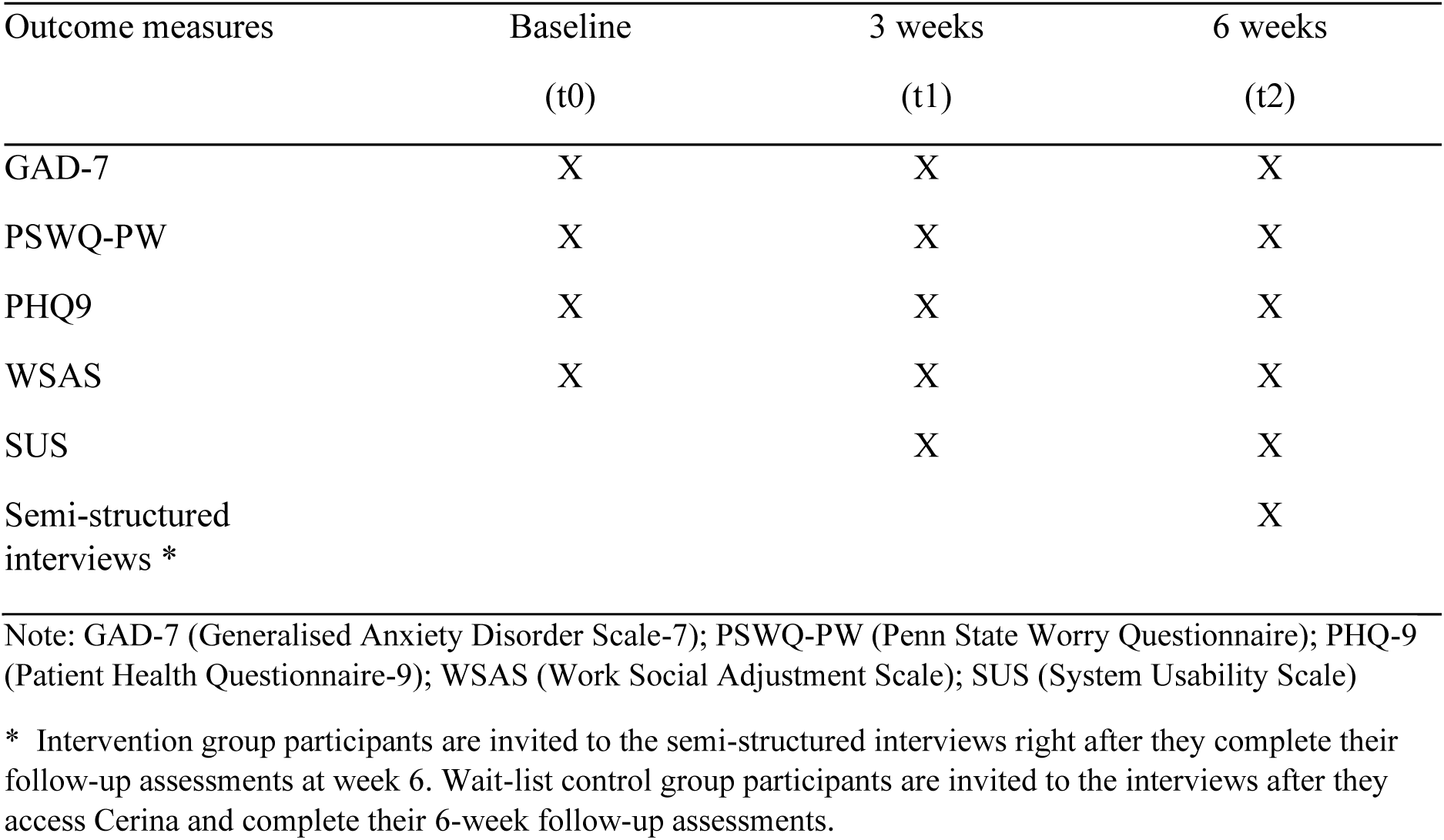
Measures collected and the time point of data collection for both groups.

### Randomization, Treatment Allocation, and Blinding

The randomization scheme will be derived using random allocation software by the Cerina technical lead who is not involved in the study. Randomization will take place in blocks of 6 and will take place in a 1:1 ratio. Allocation will be concealed from the researchers involved in the study. It is not possible to mask participants due to the waitlist control group and the nature of the intervention. All participants will be informed about the condition they are assigned to.

### Participants

Participants will be adult students attending Ulster University who have reported anxiety symptoms based on the Generalised Anxiety Disorder Scale-7 (GAD7) questionnaire.

#### Inclusion criteria

- Having mild to severe self-reported anxiety symptoms. Those who score between 5 and 19 on the GAD7 questionnaire will be accepted as eligible,
- 18 years of age and older
- Enrolled as a student at Ulster University (i.e. having a student ID number)
- Fluency in English
- Provision of an informed consent
- Have a smartphone (i.e. Android device or iPhone).
- Have an internet connection

#### Exclusion criteria

- Have minimal anxiety symptoms as defined by a score of 5 and below on the GAD7 questionnaire
- Scoring 19 and above on the GAD7 questionnaire
- Have self-reported suicidal thoughts based on their scores on the Patient Health Questionnaire-9 (PHQ9).
- Have recently (within the last 6 weeks) started taking psychotropic medication.
- Not consenting that their contact details (name, surname, email address) will be shared with the Student Wellbeing team (see the ethical and safety considerations)
- Receiving psychological treatment is not part of the exclusion criteria.

### Sample size

The sample size of the study is 90 participants (N=45 in treatment, N=45 in Wait-list control) in total. As this is a pilot RCT study and the objective is mostly to test the feasibility of the trial procedure, and intervention uptake, and evaluate the acceptability of the intervention towards building capacity for a larger test population, no formal sample size calculation was performed (46,47). The audit of the sample sizes for the pilot and feasibility RCTs indicated that the median sample size per arm across all the types of studies was 30 (48). Browne also recommended that 30 participants per condition are needed to estimate a parameter (49). Taking a possible dropout rate into account, the aim is to include 50% more participants than initially intended in both arms.

### Procedure

Recruitment will take place at the Ulster University campuses from April 2023 until April 2024.

Information about the study will be provided on the Cerina website (https://www.cerina.co/) and on the Ulster University Student Wellbeing team’s website where potential participants can register by filling out the online registration form. Those who do will receive an e-mail with an information sheet, consent form, and a link to the baseline questionnaire. Those who are eligible (based on the baseline questionnaire) and who give consent will receive another e-mail explaining which group they have been allocated to. Those who are in the treatment group will receive further instructions on how to download the Cerina app and technical assistance will be provided if needed. Those who are allocated to the wait-list control group will receive an email informing them about their group allocation and their access to the services offered by the Student Wellbeing team voluntarily while they are waiting to access Cerina.

Participants will be recruited from the Ulster University student population through several channels as described below.

The research team will liaise with the Student Wellbeing team to promote the study within their networks of students through word of mouth and employing banners and/or social media posts on Cerinàs LinkedIn and Instagram accounts, the Ulster University Student Wellbeing Facebook page, and the Ulster University Instagram account. All these communications (i.e., emails, banners, and social media posts) are prepared by the research team and reviewed by the Student Wellbeing team and the Ulster University Research Ethics Committee in terms of understandability and appropriateness of the content and the language. They will be circulated within the student email lists and/or posted on relevant social media pages.

The research team will collaborate with the Student Wellbeing team and student networks to organise online and/or face-to-face workshops to introduce the study and answer interested students’ questions.

Finally, the Ulster University Students’ Union will be approached in person and/or using e-mails to circulate information about the study within their networks.

### Data Collection Procedures

The consent forms can be signed either digitally through DocuSign or on paper. If the paper version is signed, the participants will need to email the picture and/or the scanned copy of the consent form to the research team.

After providing informed consent, participants are informed about their group status via email. All self-report assessments will be administered via email at baseline, 3-weeks, and 6-weeks follow-ups except for the System Usability Scale (SUS). SUS will only be assessed at 3 weeks and 6 weeks for intervention group participants. Wait-list group participants will complete SUS at 3 weeks and 6 weeks after their access to Cerina. Qualtrics software is used to generate all self-report assessments including Generalised Anxiety Disorder-7 (GAD7), Patient Health Questionnaire-9 (PHQ9), Penn State Worry Questionnaire (PSWQ), Work Social Adjustment Scale (WSAS) and SUS. It will take approximately 10-15 minutes to complete the self-report assessments.

Intervention group participants will be invited to the semi-structured interviews once they complete their 6-week follow-up assessments. Wait-list group participants will be invited to these interviews after accessing Cerina, and once they complete their 6-week follow-up assessments. The data collection for the semi-structured interviews will last approximately 40 minutes, will be remote (online via the Microsoft Teams account of Ulster University), and will be facilitated by the members of the research team. The interviews will be digitally (audio and video) recorded and transcribed verbatim based on participants’ consent. Participants will be able to opt out of the video recording or they could choose to complete an online post-assessment feedback questionnaire (or neither) if they wish to give more in-depth feedback on the usability aspects and the clinical content of Cerina. The online post-assessment feedback questionnaire is generated through Qualtrics software (same as other self-report questionnaires used in the study) and includes the same questions as the topic guide. The response format is open-ended to give participants more space to express themselves.

### Patient Public Involvement Statement

We have used the NIHR INCLUDE framework (50) to ensure the project considers equality, diversity, and inclusivity in delivering its objectives. During the product development phase, we ran a series of workshops and interviews with key stakeholders (aged 18-34, N=4) including a product expert, researcher (i.e. with expertise in digital interventions), policy expert (i.e. with expertise in policies regarding data protection, safety and implementation) and a university student between October 2022 and April 2023. The group highlighted the key priorities to facilitate engagement with the intended users such as customised notifications, chatbots and flexibility to revisit the completed sessions and exercises whenever they want. Their feedback and the usability and/or data protection-related concerns were discussed with Cerinàs technical team and the clinical safety officer. As a result, decisions were made regarding the User Interface and the engagement features. This co-design process involved identifying intended users’ needs and preferences based on the literature and the previous users’ feedback on Cerina, defining intervention features to address the user needs and preferences, designing content and visual features and testing prototypes in accordance with the recommended guidelines (51). Design and intervention features were incorporated into the final intervention as described below. Outcomes from the codesign workshops and interviews were disseminated to the group via email. The potential burden of the intervention and possible bugs, glitches and/or typos were also assessed by the group and amendments were made accordingly. The group will not be involved in the recruitment and/or conduct of the study.

The study will be conducted in partnership with Ulster University. Additionally, the research team has a good working relationship with the Ulster University Student Wellbeing Team. During the design stage of the study, the Ulster University Student Wellbeing team was engaged extensively. They reviewed the recruitment materials and they have been involved in the key decisions regarding the signposting and risk assessment procedures. The research team will work closely with the Ulster University Student Wellbeing team during the recruitment process to ensure that students will go through further assessment with advisors and they will be signposted to the available services whenever needed. Additionally, the partnership with Ulster University will also ensure the reach and access to Ulster University students from across different campuses.

Finally, all the involved parties (Ulster University, Ulster University Student Wellbeing team, IT4Anxiety assessment and validation team) will be involved in reviewing the recruitment and signposting processes and dissemination of the findings.

### Intervention

The intervention is unguided, therefore it does not provide any professional support related to the therapeutic content of the sessions (52). If participants experience any difficulty with a particular therapeutic technique on the app and/or a technical problem, they are advised to contact the Cerina team at. If the users experience a distressing situation unrelated to Cerina, they are advised to contact their General Practitioner and/or a health care professional. Also, there is a self-care page available on the app including links to external resources for further help such as National Health Services (NHS) helpline, mental health charities, support groups and Samaritans.

Cerina consists of 7 sessions of CBT for the treatment of GAD. Each session contains a range of information and tasks/exercises to help the user understand the condition of GAD, the treatment approach, and how it will apply to them. The intervention is based on an evidence-based treatment protocol; hence the sessions will flow from one to the other and the user will complete the sessions in a progressive direction. Participants are advised to complete the first six sessions on a weekly basis. However, they can repeat a session before going on to the next session. The last session is designed as a review of progress session rather than a full therapy session including exercises and homework assignments. Once all sessions have been completed, participants can go back over any of the sessions. There are anxiety management exercises, which the user can go to whenever they wish. There are also a therapy reflection journal and self-care resources including further anxiety management techniques, resources, and podcasts that the user will have access to whenever they want.

A summary of the content of each session is as follows:

1. **Learning About GAD:** The first session provides the user with psycho-education as an introduction to GAD as a mental health condition and will ask them to consider whether they recognise the features in themselves.
2. **Dealing with worry** – using the worry tree: The aim of this session is to provide the user with psycho-education on the types of worries. The user starts conceptualising their worries and making decisions about how to manage different types of worries through the technique called “worry tree”.
3. **Managing worry and anxiety**: In this session, the user learns about the link between worry and anxiety and they gain a deeper understanding of the vicious cycle (i.e. particular situations trigger worry thoughts and they lead to physiological and psychological reactions and noticing those anxious symptoms lead to more worrying). The worry management techniques such as worry postponement, and attention focus training, are also introduced to help the user break this vicious cycle.
4. **Beliefs about worry**: This session introduces core beliefs about worrying which perpetuate the worry thoughts. The user learns about the advantages and disadvantages of their beliefs and how helpful they are in their life.
5. **Managing uncertainty**: This session helps the user manage uncertainty through problem-solving and imaginal exposure techniques. The user learns to deal with their worries and develops an action plan in a step-by-step manner through problem-solving techniques. The latter helps them to spend less time and energy on their worries gradually through imaginal exposure.
6. **Testing beliefs:** This session aims to guide the user to test their beliefs through behavioural experiments. They also learn to do a cost-benefit analysis of worrying to help them order their thinking and be more objective.
7. **Review and therapy blueprint**: This session helps the user to review their goals and what they have learned during the therapy, and to think about how they will take this learning forward to maintain their progress in continuing to manage their GAD symptoms in the future.

Design features identified in the patient and public involvement phase were included in the intervention to promote user engagement. These include (1) vibrant and simple illustrations to present the case studies in each sessions; (2) customised notification settings; (3) a separate self-care page including pod casts and available resources for further help if needed; (4) regular check-ins to rate mood; (5) embedding a safety mechanism within the user’s conversations with the chatbot. For instance, if a user talks about death and suicidal thoughts, the chatbot provides contact details of the available crisis lines and services for further help; and (6) providing more flexibility regarding the navigation. For instance, the users are able to start where they are left off once they login to the app, and they can go back and review their therapy journey whenever they want.

### Wait-list Control Condition

Participants in the waiting list control condition will have access to the on-campus wellbeing services offered by the Student Wellbeing team should they wish to use the services. The Student Wellbeing team on the Belfast, Coleraine, and Magee campuses provides free and confidential support and guidance to students with a broad range of issues, concerns, and challenges, helping them to successfully engage in their studies. Student Wellbeing Assistants provide an initial assessment to determine a student’s primary need and then Wellbeing Advisers, Student Mental Health Advisers, and Accessibility Advisers are available to provide a variety of solution-focused interventions based on that need. The Accessibility Advisers also help students with disabilities including diagnosed mental health conditions to get additional support and access funded disability support through Disabled Students’ Allowance. Each campus team is led by an experienced Student Wellbeing Manager to support the advisers in the management of risk and lead to an efficient and robust response if a Clinical Incident occurs. Additionally, therapeutic counselling support is also available free to students through the external counselling provider via a dedicated 24/7 counselling helpline. The Student Wellbeing Team works closely with the counselling provider to monitor student engagement and promote the service as appropriate (For further information, please see https://www.ulster.ac.uk/wellbeing/health-and-wellbeing).

## Materials

### Primary outcome measure

1. Feasibility is defined as adherence to the intervention, its usability and potential to deliver a full trial in the future (53,54). Adherence is defined based Staudt’s model of adherence (ref) and it includes behavioural and attitudinal adherence (55,56). Behavioural adherence refers to the participant’s performance on the tasks that are necessary to implement an intervention in daily life such as, homework completion (55,56). Attitudinal adherence refers to the emotional investment and commitment to the intervention (55,56). To measure behavioural adherence to Cerina, we will collect data on the number (%) of the login attempts of the participants per week and per month, the number (%) of intervention group participants with at least 1 session attended, and the number (%) of intervention group participants with desired number of sessions (N=6) completed. Furthermore, to measure attitudinal adherence, we will collect data on the number (%) of the therapeutic content completed by the participants. We will collect data on whether participants complete the core therapeutic content (i.e. have entries in their worry diary in session 1 and reflect on their worry diary in session 2), some therapeutic content (i.e. psychoeducational part) or no therapeutic content at all (i.e. few taps, invalid entries). The usability of Cerina will be measured with the System Usability Scale (SUS) (57) and with semi-structured interviews. An example item: “I think I would use Cerina frequently”. The SUS is composed of 10 statements that are scored on a 5-point scale of the extent of agreement (score 0 to 100). The reliability is good (Cronbach’s alpha 0.91) (57). Interventions with scores of 70 and above are accepted as highly usable (57) and scores between 50 and 70 indicate acceptable usability of an intervention. Interventions with scores of 50 and below are subject to concerns about their usability by the target population and should be investigated further (57). In order to investigate the potential to deliver a full scale RCT in the future, we will assess the response rates to the self-report questionnaires, examine the recruitment procedures and participants’ willingness to be randomized to a waiting list control group (53).

### Secondary outcome measures

2. The Generalised Anxiety Disorder Scale-7 (GAD-7) (58) is a 7-item self-report scale that identifies and measures the severity of GAD. An example item for GAD7: “Worrying too much about different things”. Scores range from 0 to 21, with a cut-off score of 5 distinguishing between clinical and non-clinical populations. The scale has good psychometric properties (58).
3. The Penn State Worry Questionnaire (PSWQ-PW) is a 15-item inventory assessing both the weekly status of pathological worry and treatment-related changes of worry during the treatment (59). An example item: “If I didn’t have enough time to do everything, I didn’t worry about it”. Each item is scored on a 7-point rating scale, ranging from never 0 (never) to 6 (almost always). The total score ranges from 0 to 90 with a high score indicating more worrying. PSWQ-PW shows good reliability and convergent validity (59).
4. The Patient Health Questionnaire (PHQ-9) (42) is a 9-item self-report scale that measures depression symptoms. An example item: “Little interest or pleasure in doing things”. Scores range from 0 to 27, with a score of 10 and above considered to be a clinically significant level of depression. The PHQ-9 has good reliability and validity (60).
5. The Work and Social Adjustment Scale (WSAS) (61) is a 5 item self-report measure that assesses functional impairment. An example item: “Because of my mental health, my ability to work is impaired”. Scores range from 0 to 40. The scale assesses the impact on work, home, social and private activities, and personal and family relationships. A score of 20 and above is considered to indicate severe functional impairment, scores between 10 and 20 suggest severe but functional impairment and scores of 10 and less are considered subclinical. The scale has good reliability and validity (61).

### Process evaluation

Semi-structured interviews will be conducted with the participants who complete the intervention and consented to be interviewed. The topic guide for the interview was developed by the research team based on the Consolidated Framework for Implementation Research (CFIR) (62). The core topics included participants’ overall experience and perceptions on the usability of the UI of Cerina, existing pathways and barriers to using the app in daily life, the clinical utility of the application, the contexts in which the implementation of the clinical content takes place and the processes of intervention delivery. We will recruit until data saturation (63), with an estimated sample size of 20 participants. Descriptions of the context of each interview will be documented in fieldnotes to support analysis.

Draft interview schedules have been developed and amended by the research team based on the CFIR framework and feedback from the participants from the previous studies. Additionally, they have been reviewed by the Ulster Student Wellbeing team. The topic guide focuses on:

- The usability of the intervention (i.e. potential barriers and facilitators whilst using the Cerina app in daily life)
- Whether the participants relate to the User Interface and/or content of the intervention
- How the clinical content of the intervention help participants understand and manage their anxiety symptoms and which specific exercises help and/or do not help them while they are using the intervention.

### Data analysis plan

The recruitment and consent rates will be carefully monitored according to the CONSORT guidelines. The group allocation procedure will be monitored by the technical lead of Cerina (independent researcher). The response rates to the study questionnaires and the adherence rates will be monitored by the corresponding author.

The process evaluation will be investigated through semi-structured interviews. These interviews will be audio-recorded and transcribed. The transcripts will be analysed thematically and independently by two researchers (one experienced qualitative researcher and one researcher who is involved in the data collection) in NVivo software. The deductive coding will be informed by the CFIR framework (62). Additionally, there will be an inductive coding. Coders will meet each other regularly and the wider study team to discuss codes and begin by identifying and indexing themes relevant to an interview (64). Also, they will explore relationships between descriptive themes (63). The identified themes will be interrogated about each account, as a means of understanding a particular case; compared across cases by highlighting potential similarities and differences between respondents; and, finally, related to those characteristics of the respondent that could be reasonably justified as an explanation which mediated experience (65).

The preliminary effects of Cerina in reducing elevated levels of anxiety and worrying among students will be tested through the linear mixed models in RStudio version 3.6.1 (66). We will compare reductions in primary/secondary outcomes between and within groups across two time points in the intention-to-treat sample, we will use linear mixed models in RStudio version 3.6.1 (66). This method allows the number of observations to vary between participants and handles missing outcome data.

The mixed model uses a longitudinal data structure that includes both fixed and random effects. Time (categorical), group (treatment *v*. wait-list control), and interactions between group and time will be included as fixed effects in mixed models together with a random intercept and random time effect. Differences in least-squares mean (intervention effects) at each time point with 95% confidence intervals will be derived. Cohen’s *d* for the effect of the intervention will be estimated by calculating the difference between estimated means (corrected for baseline) divided by raw pooled standard deviation. Effect sizes of 0.8 are accepted as large, effect sizes of 0.5 are moderate and effect sizes of 0.2 are small (67). A two-sided *p* < 0.05 indicates statistical significance. These analyses will be done based on the intention-to-treat sample and missing data will be handled through multiple imputations.

The reliable change index will also be used to evaluate whether participants have reliable and clinically significant change scores from baseline to post-assessment (68).

### Ethics and Dissemination

Ethical approval for the study has been granted by the Ulster University Research Ethics Committee (ID: FCPSY-22-084). Cerina includes information on crisis lines in the content of the application.

Participants can go to a page where further information on self-care resources and help-seeking are provided within the application. Additionally, receiving psychological treatment is not exclusion criteria and all participants are signposted to the services offered by the Student Wellbeing team, their GP, and/or additional resources (e.g. crisis lines, Lifeline etc) and are encouraged to use these services regularly through online communication throughout their participation. Eligible participants also need to agree on sharing their student ID numbers with the Student Wellbeing team. This is necessary for two main reasons: 1) to signpost participants to the Student Wellbeing when needed and, 2) to be able to monitor the use of additional psychological services among all participants throughout the study.

Interested participants who are currently on psychotropic medication or presenting minimal and/or severe anxiety symptoms (as measured by the GAD7) and/or reporting suicidal thoughts during the baseline assessment will receive an e-mail about their results and will be informed that Cerina is not designed to meet their needs and they will be signposted to the Student Wellbeing team for further help. In that email, the crisis lines and resources for further help will be provided.

Another potential risk is that some participants might get worse whilst they are using Cerina. The existing studies testing CBT-based mobile applications in treating GAD symptoms indicate that this is a rare risk (19,47). We will monitor this risk based on participants’ responses to the online questionnaires. We will also monitor their answers to the regular check-in questions asking about their mood whenever they log in to the app. If participants’ scores are getting worse and/or their check-in questions indicate that they might be feeling low lately, they will receive a notification on the app directing them to the self-care page, anxiety management exercises, and/or the resources pages for further help. They will also receive an option as a notification, to be referred to the Student Wellbeing team for further risk assessment. If they choose this option, the research team will notify the Student Wellbeing team to schedule a meeting with them for risk assessment and further help.

The results of the study will be disseminated through publications in scientific journals, presentations at relevant congresses, conferences and/or public events.

### Findings to Date

As of 31 March 2024, we have recruited 158 (N=79 intervention; N=79 wait-list control group) participants. In the intervention group, a total of 29 participants completed their 3-week assessments, and a total of 47 participants completed their 6-week post-assessments. In the wait-list control group, a total of 43 participants completed their 3-week assessments, and a total of 56 participants completed their 6-week assessments. Additionally, we have conducted 4 interviews with the participants and a total of 16 participants opted out of the interviews but completed post-assessment feedback questionnaires.

## Discussion

The current paper describes a study protocol of a pilot feasibility RCT with two main objectives: to learn about the feasibility (i.e. adherence to the intervention, its usability and potential to deliver a full trial in the future) of delivering Cerina among a student population; and 2) to test the preliminary effects of Cerina in reducing GAD symptoms compared to a wait-list control group among Ulster University students presenting with self-reported GAD symptoms.

One of the strengths of this study is that it proposes to test a low-threshold CBT-based mobile application among Ulster University student population who have low receipt of on-campus and external psychological support services despite their vulnerability for mental health problems (23). Digital CBT based interventions aims to provide easy access, anonymity, reduced costs and stigma attached to help-seeking (30). However, there are mixed results for the effectiveness of these interventions among student populations (31–37). Additionally, recent studies point to low engagement with CBT-based mobile applications in particular among student populations despite the assumed benefits (42). Ciharova and colleagues (69) investigated the reasons for drop-out from the ICare Prevent intervention among university students in the Netherlands (69). There were several person- and intervention-related reasons for dropout most commonly, lengthy sessions colliding with students’ busy academic schedules, needs for different kinds of help and/or absence of personal contact (69). Although Cerina is an unguided intervention, we expect that the design features such as customised notification settings, flexibility in navigation and the chatbot function will address some of the key person- and intervention-related drop-out reasons identified by Ciharova and colleagues (69). Therefore, this study will provide an opportunity to contribute to the growing evidence base on the role of emerging technology-based engagement features, and their impact on the effectiveness of digital CBT-based interventions.

Another strength of this study is that it is based on a partnership and a collaboration between Ulster University, Ulster University Student Wellbeing team and Cerina, a mental health start-up. All these parties have been involved in making key decisions about the recruitment strategies, risk assessment and signposting procedures as well as dissemination plans. A recent study by Lattie and colleagues (42) highlighted the digital overload among university students during and post-pandemic and the ineffectiveness of email and social media communication strategies during the recruitment process. Additionally, they note the disruption caused by the pandemic had a negative impact on the communications between the research team and the student wellbeing staff as the staff members were overburdened by competing priorities (42). In the current study, we expect that the established partnership and the utilisation of various recruitment channels will help reach and access to the intended population, and such challenges specific to the pandemic context will not pose problems.

The limitations of the current study must also be acknowledged. First of all, the study is designed as a feasibility trial and it is not powered to detect statistically significant effects of the intervention (46,47). Second, although the study participants will be recruited from across the four campuses of Ulster University, the results might not generalise beyond the inclusion and exclusion criteria of the current study. Third, the wait-list control group has been chosen as a comparison group in the current study to offer the possibility of completing the intervention to a vulnerable population who might not have access to any form of psychological help otherwise. It is known that wait-list control groups motivate people to participate in the control group and the trial, and lead to high expectations of the effects of the intervention (70). It is also known that high expectations result in better outcomes of interventions. Therefore, the wait-list control group may considerably overestimate the effects of an intervention in the current study (70).

To conclude, this study has several strengths which will contribute to the literature about digital CBT-based interventions for treating generalised anxiety symptoms among university students. These are established partnerships with the relevant stakeholders strengthening the reach and access to the intended population, engagement features which are incorporated into the Cerina intervention based on the co-design principles, and low-intensity CBT-based mobile application targeting Ulster University students.

## Data Availability

All data produced in the present study are available upon reasonable request to the authors

## Authors’ contributions

OEB had the idea for the study, and OEB and GL designed the study. SP, TR, PM, and JC contributed to the conception of the work. MI wrote the treatment protocol for the intervention. KA and TR contributed to the literature search. All authors read and contributed to the revisions and the final paper.

## Acknowledgements

The authors would like to acknowledge Omkar Kadam (Cerina Head of Innovation), Priyadharshini Rajaram (Cerina Technical Product Manager) and Charlotte Monnickendam (Cerina Head of Partnerships) for their work and dedication to product development and innovation. They also wish to thank the Cerina Board of Directors Andrew Vallance Owen, Anoop Aggarwal and the rest of the advisors for their continuous support for Cerinàs evidence-based approach. Finally, the authors would like to thank the stakeholder group who is involved in the product development.

